# Optimizing school meals in Ghana: integrating new food and nutrient standards with aspects of affordability, cultural acceptability and environmental sustainability

**DOI:** 10.1101/2025.06.17.25329697

**Authors:** Patricia Eustachio Colombo, Alexandr Parlesak, Gloria Folson, Boateng Bannerman, Audrey Anang-Tetteh, Gabriel Ador, Lois Ackah-Swanzy, Melissa Vargas, Richmond Aryeetey, Aulo Gelli

**Affiliations:** Centre on Climate Change and Planetary Health, London School of Hygiene and Tropical Medicine, Keppel st, London WC1E 7HT, UK; Department of Medicine, Karolinska Institutet, 141 83 Huddinge, Sweden; Institute of Nutrition, Exercise, and Sports (NEXS), University of Copenhagen, Copenhagen, Denmark; Personalized Nutrition, Baden-Wuerttemberg Cooperative State University (DHBW), Heilbronn, Germany; Department of Nutrition, Noguchi Memorial Institute for Medical Research, University of Ghana, Legon, Ghana; Food and Nutrition Division, Food and Agriculture Organization of the United Nations, Rome, Italy; Division of Human Nutrition and Health, Wageningen University and Research, Wageningen, The Netherlands; School of Public Health, University of Ghana, Legon, Accra, Ghana; International Food Policy Research Institute (IFPRI), Washington DC, USA

**Author notes:** Correspondence: Dr Patricia Eustachio Colombo, Department of Medicine, Karolinska Institutet, 141 83 Huddinge, Sweden, Assistant professor.

**Keywords:** child nutrition, food security, food sustainability, public procurement, dietary guidelines

## Abstract

**Objectives:** To optimize school food baskets in Ghana to meet newly proposed food and nutrition targets while considering cultural acceptability and cost.

**Design:** Modelling study. Data on existing school meal menus was collected from various regions to provide baseline inputs. Linear programming was used to model school meal baskets that satisfied minimum nutrient and food targets for school meals while meeting cost and acceptability constrains. The study included five optimization scenarios, each varying in budget constraints and acceptability/food-based parameters.

**Setting:** Ghana

**Participants:** NA

**Results:** Baseline school food baskets were significantly deficient in energy, protein, and essential micronutrients compared to food and nutrient standards for school meals in Ghana. Optimization resulted in school food baskets that met cost, nutrient and food-based/acceptability targets but with substantial deviations from baseline. It also resulted in increased reliance on animal products and higher environmental impacts, highlighting trade-offs with environmental sustainability.

**Conclusion:** The study underscores LP’s potential for enhancing school meals in Ghana but highlights the need for increased financial investment for reaching dietary goals. Addressing local realities and cultural preferences is essential for implementing effective, sustainable school meal strategies and improving child health.

## Introduction

Undernutrition is estimated to affect over 700 million people globally (1). Childhood malnutrition has long-term consequences for development and health (2) and deficiencies in micronutrients impair children’s physical and psychosocial health and development (3). Addressing health and nutrition risks throughout childhood and adolescence is critical to ensure optimal development and to protect earlier life course investments (4). School feeding programs are popular safety nets that reach over 400 million children every school day (5). The evidence on school feeding programs indicates a variety of potential contributions to children’s education, health, nutrition, and social protection (6).

In Ghana, where food insecurity and malnutrition remain pressing challenges (7), school meals have been recognized as a key strategy to enhance both the health and educational outcomes of children. Currently reaching over 3.8 million primary school children, the Ghana School Feeding Program (GSFP) is the country’s largest safety net (8). The GSFP is a multisectoral program with objectives across social protection, education, health, nutrition and agriculture dimensions. A rigorous impact evaluation of the GSFP at scale indicated that the program can improve cognition and learning outcomes (9). The program has not demonstrated improvements to height-for-age and BMI-for-age z scores overall, however, positive effects on height-for-age have been observed in 5-8y old children, girls and in children from poorer households (10). The GSFP menus based on a 0.12 USD per child budget are designed to meet approximately 30% of recommended daily energy and nutrient requirements of school children, and include foods grown by smallholder farmers (11).

However, persistent gaps in financing, food quantity, and food quality (12), hinder the successful realization of these goals. Furthermore, little is known about the environmental impacts of school meals in Ghana, which is an important knowledge gap given the growing emphasis on sustainability in food systems.

A supply chain study of the GSFP reported that the main challenges faced by caterers providing the meals included the inability to mitigate the effects rapid changes in food prices (13). Caterers respond to these challenges frequently by adapting menus, i.e., by reducing portion sizes or by adjusting the quality of the food. The rising costs of food and increasing economic pressure necessitate a re-evaluation of existing meal plans to ensure that they remain financially sustainable while responding to Ghanaian children’s critical dietary gaps.

Linear programming (LP) has been demonstrated to be an effective tool in designing cost-effective, nutritionally adequate diets for Ghanaian families (14). The method has also successfully been applied in school canteen settings (15–17). This study uses LP,, to optimize school food baskets to meet newly proposed food and nutrient targets for school meals based on Ghanaian children’s actual dietary gaps, while considering critical factors such as food affordability, operational costs, anticipated inflation, cultural acceptability, and environmental impacts.

## Material and methods

### Study design

This was a modelling study using established algorithms of linear programming (18) to design school food baskets that fulfilled newly developed food and nutrient targets for school meals in Ghana (19). This study considered aspects related to food affordability (food costs, non-food costs, anticipated inflation), cultural acceptability (minimum deviation from reported baseline usage of foods) and environmental impacts (greenhouse gas emissions and water use). The optimization analysis was performed on two distinct school food baskets. First, we optimized the current school food basket. Second, we optimized an improved food basket, which consisted of the current recipes modified by a nutritionist to enhance protein content and quality (see more details below).

### Baseline school food baskets

#### Base menus

Data on current school meal menus, including recipes and ingredients, were collected in ten districts (Ada East, Akyenmansa, Amansie South, Atwima Mponua, Bongo, Nanumba South, Nkwanta North, South Tongu, Tain, and Wa West) across 5 regions in Ghana. Schools were selected by the GSFP on the basis of having a functioning program. These menus were collected by a team of 20 trained enumerators from the Noguchi Memorial Institute for Medical Research at the University of Ghana. Data collection took place in September 2023, following standard operating procedures (SOPs). Briefly, at the beginning of each day, the teams of enumerators would visit the targeted schools and begin the calibration of the different measurement scales. Weights of cooking pots used for the meal preparation were recorded, alongside the measurement of weights of raw ingredients before and after they had been cleaned and prepared for cooking. The weight of the cooked meals was then recorded after cooking to allow for the calculation of ingredient proportions and recipes for mixed dishes using the simple ingredient method (20).

These collected menus (i.e. “Base menus”) (see dishes and recipes in Supplementary Methods) were aggregated and averaged; each individual food item was averaged by dividing its total amount by the number of recipes (where each recipe was intended for one child, consumed once per day) to calculate the average amount (in grams) per child and day. These average amounts of individual food items per child and day were used to construct the first baseline school food basket, upon which the first set of models were applied using LP (Models 1-2, Table 1). The baseline school food basket for the Base menus contained a total of 45 individual food items together with corresponding amounts used (g/child/day) for cooking, and average prices (USD/kg of food). Price information was collected from caterers based on their actual purchases. Additional price information was obtained from the price monitoring included in the School Meal Planner Plus database (21).

**Table 1.** Characteristics of all applied models.

| Model nr | Objective function (minimum) | Decision variables | Nutritional constraints | Cost constraints | Food based recommendations<sup>e</sup> | Acceptability constraints |
| --- | --- | --- | --- | --- | --- | --- |
| 1 | | | | \$0.12 | | |
| 2 | | | | \$0.10<sup>c</sup>; food prices subjected to 12% increase due to anticipated inflation | | |
| 3 | TRD<sup>a</sup> from baseline food supply | Amount of individual foods | Meet all nutrient targets<sup>b</sup> | \$0.10<sup>d</sup>; food prices subjected to 12% increase due to anticipated inflation | Daily intake of:<br>≥133g fruit and vegetables,<br>≥30g legumes nuts and seeds, and<br>≥20g animal protein | Min RD<sup>f</sup> for food items set to 75-80%; keep at least 75% of original grain amount |
| 4 | | | | \$0.13<sup>d</sup>; food prices subjected to 12% increase due to anticipated inflation | | |
| 5 |  |  |  | No cost constraint, food prices subjected to 12% increase due to anticipated inflation |  |  |

<sup>a</sup>Total Relative Deviation.
<sup>b</sup>As per the Dietary Reference Values for school meals in Ghana.
<sup>c</sup>Budget available when subtracting 20% operational cost from \$0.12.
<sup>d</sup> Budget available when subtracting 20% operational cost from \$0.16.
<sup>e</sup>As per food targets and mealtime recommendations for school meals (lunch) in Ghana (19).
<sup>f</sup>Relative Deviation from baseline food consumption.

#### Improved menus

A set of manual modifications were made on the observed recipes by a senior nutritionist at the Noguchi Memorial Institute for Medical Research with a goal of improving the protein quality and total protein content of school meals, including the addition of eggs, texturized soy protein and other protein-rich foods, factoring for an additional budget allowance of US$ 0.16. This was based on costs for the additional protein that was going to be provided.

These modified menus (i.e. “Improved menus”) (see dishes and recipes in Supplementary Methods) were aggregated and averaged; each individual food item was averaged by dividing its total amount by the number of recipes (where each recipe was intended for one child, consumed once per day) to calculate the average amount (in grams) per child and day. These average amounts of individual food items per child and day were used to construct the second baseline school food basket, upon which the second set of models were applied using LP (Models 3-5, Table 1). The baseline school food basket for the Improved menus contained a total of 29 individual food items together with corresponding amounts used (g/child/day) for cooking, and average prices (USD/kg of food).

#### Nutrient content and environmental impacts

The gram quantities of various food items in the baseline school food baskets for both the Base and Improved menus were linked to their respective nutrient composition using the West African food-composition table and the RING nutrient composition table, which is a compilation of food-composition databases relevant to Ghana (22). We also calculated greenhouse gas emissions (GHGE) and water use for both baseline and optimized menus using values adapted from Poore & Nemecek’s systematic review of global environmental footprints (23). GHGE were aggregated into global warming potential in carbon dioxide equivalents (CO_2_eq) using the International Panel on Climate Change (2013) characterization factors with climate-carbon feedbacks. Water use (WU) was determined based on the freshwater withdrawals associated with food consumption, encompassing irrigation water, water for animals to drink, and water utilized in food processing (23). To accurately assess the impact of each food item, we incorporated information on whether the food was imported or locally produced in Ghana (23).

### Optimization

Linear programming (LP) is a powerful tool that has been effectively employed to optimize dietary objectives while navigating a variety of sometimes conflicting constraints (24,25). At its core, this method involves applying an algorithm to either maximize or minimize a linear objective function, which represents the variable of interest. This optimization is bound by a set of linear constraints, which are predefined requirements that must be met in the context of decision variables— specifically, the quantity of each food item included in the menus (26). A solution is deemed feasible when all specified constraints are satisfied. However, if the selected constraints are excessively strict, the algorithm may fail to yield a solution, indicating that there is no feasible answer to the associated mathematical problem. The constraints that are critical in determining whether the objective function can be maximized or minimized—those fully satisfied within their established limits—are referred to as “active constraints” (27). For this analysis, linear optimization was carried out using the COIN-OR Branch and Cut Solver algorithm, which is part of the OpenSolver add-in for Excel® 2016, version 2.9.0 (28).

We optimized school food baskets in Ghana according to the five different models (Table 1). The relative deviation (RD) from the average amount of each food item was calculated as RD = (w_opt_ – w_bas_)/w_bas_, where w_opt_ is the food weight in the optimized school food basket and w_bas_ is the baseline food basket. As the objective function of all LP models, we chose the minimization of the total relative deviation (TRD, equaling the sum of RDs for all foods considered) from the baseline food basket (18,29). This objective function was implemented to maximize the similarity between the baseline and the optimized food baskets. The decision variables were the amounts of individual food items in the different models. To implement the absolute values of RD into the linear programming process, the mathematical technique of Darmon and colleagues (18) was applied.

All optimizations applied newly developed (albeit preliminary) nutrient and food targets for school meals in Ghana as obligatory constraints. As for energy and macro/micronutrients, this meant implementing minimum thresholds for kcal, protein, iron, zinc, vitamin A, vitamin C, folate and vitamin B12 (Table 2) as per the newly developed food and nutrient targets for school meals in Ghana (19). The food targets per meal comprised the minimum amount of 133g of fruit and vegetables, 30g of legumes, nuts and seeds, and at least 20g of animal source foods (beef, fish, eggs). These targets were set specifically for the GSFP by a working group convened by the Food and Agriculture Organization of the United Nations (FAO) following a new FAO-World Food Programme methodology for this purpose (19,30), which considers contextual dietary gaps in children’s diets. In cases where the targets differed depending on age category, the nutritional constraints were averaged (i.e. the average of all age groups was used).

**Table 2.** Nutrient coverage of baseline and optimised school food supplies.

|  | Nutrient target<br>for a school<br>lunch | Baseline<br>Base<br>menus<sup>a</sup> | Model 1 | Mode<br>1 2 | Baseline<br>Improve<br>d menus<sup>b</sup> | Mode<br>1 3 | Mode<br>1 4 | Mode<br>1 5 |
| --- | --- | --- | --- | --- | --- | --- | --- | --- |
|  | % coverage of nutrient target |  |  |  |  |  |  |  |
| Energy (kcal) | ≥450 | 28 | 100 | 100 | 54 | 100 | 100 | 102 |
| Protein (g) | ≥7.3 | 41 | 145 | 158 | 120 | 450 | 423 | 301 |
| Iron (mg) | ≥2.8 | 45 | 139 | 183 | 174 | 155 | 175 | 276 |
| Zinc (mg) | ≥1.9 | 29 | 100 | 100 | 69 | 159 | 159 | 165 |
| VitA_RAE (µg) | ≥162 | 47 | 100 | 100 | 77 | 100 | 100 | 196 |
| Folate (µg) | ≥49 | 85 | 311 | 389 | 104 | 263 | 249 | 326 |
| VitaminB12 (µg) | ≥0.67 | 28 | 100 | 115 | 16 | 402 | 366 | 100 |
| VitaminC (mg) | ≥17 | 16 | 213 | 100 | 34 | 100 | 100 | 124 |

<sup>a</sup>Baseline food supply for models 1-2.
<sup>b</sup>Baseline food supply for models 3-5.

Cost constraints were implemented in all models (Table 1). Our initial model (Model 1) was constrained to cost equal to or less than the *current* budget of ∼GHC1.5 ($0.12)/child/meal. This budget is in theory meant to cover both food and non-food (i.e. operational) costs incurred by caterers. However, there are currently no guidelines or data on how that budget should be/is split. Our fieldwork in the abovementioned districts suggested that approximately 20% of funds for school meals are used for operational costs (such as salaries for cooks, electricity). Hence, the budget in Model 2 was $0.10 ($0.12 minus 20%). Since food prices are steadily increasing due to inflation, we also implemented a 12% price increase to all foods in the school food baskets (representing inflation trajectory projected for 3 months into the future) in Model 2. Model 3, based on improved recipes, had the same budget as Model 2, i.e. the current budget, and did also consider operational costs and inflation. Model 4, also based on improved recipes, had a slightly higher budget constraint of $0.13 ($0.16 minus 20%) while also compensating for the assumed cost effects of inflation. This higher budget has been suggested by stakeholders to allow for more diverse and nutritious school meals. The last model (Model 5, also based on the improved recipes) had no budget constraint to allow us to explore what a nutritious and culturally acceptable school food basket would look like if no cost limits existed. This particular scenario was aimed at informing current school meal policy and financing developments for the GSFP.

For all models/scenarios, individual food item quantities were allowed to increase unlimitedly relative to their respective baseline weights; however, similarly to previous optimisation studies (15,16,29), we limited individual food quantities to be reduced by a maximum up to 80% as an acceptability constraint (Table 1). The average relative deviation (ARD) from baseline food basket (i.e. the TRD divided by the total number of food items included in the model) was calculated as an output and used as a measure of similarity between the baseline and the optimized food basket and as an assumed proxy for cultural acceptability. Active nutrient constraints (those meeting exactly 100% of the applied limit (27)) were identified for each solution.

Climate (g CO_2_eq/child/meal) and water footprints (liters/child/meal) of the baseline and optimized food baskets were calculated for exploratory purposes but were not constrained in the models because of lack of national targets on these parameters.

In summary, five scenarios/models were explored using linear programming (Table 1) to attain school food baskets that fulfilled all nutrient constraints and were as similar as possible to:

1. the *current* school food basket, cost equal to or less than the *current budget* of ∼GHC1.5 ($0.12)/child/meal, and considering cultural acceptability and food-based constraints if possible
2. the *current* school food basket, cost equal to or less than the *current budget* of ∼GHC1.5 ($0.10)/child/meal, accounting for operational costs and inflation, and considering cultural acceptability and food-based constraints if possible
3. an *improved* school food basket, cost equal to or less than the *current budget* of ∼GHC1.5 ($0.10)/child/meal accounting for operational costs and inflation, and considering cultural acceptability and food-based constraints if possible
4. an *improved* school food basket and cost equal to or less than an *enhanced budget* of *∼GHC2* ($0.13)/child/meal accounting for operational costs and inflation, and considering cultural acceptability and food-based constraints if possible
5. an *improved* school food basket *without a budget constraint* accounting for operational costs and inflation, and considering cultural acceptability and food-based constraints if possible

We adopted an iterative approach where we aimed to apply all constraints (Table 1) if possible, and where we had the following order of priorities as for achieving the suggested constraints: 1) budget and nutrient constraints, 2) acceptability constraints; 3) food-based constraints.

## Results

### Baseline school food baskets

The baseline school food basket for the Base menus provided less energy and protein than the defined school meal nutrient targets (Table 2). It also fell short of the targets for all micronutrients. The baseline school food basket for the Improved menus was slightly less deficient; this food basket was lower than recommended with respect to the targets for energy, zinc, and vitamins A, B12 and C. The cost of the baseline school food baskets was US$ 0.10 (Base menus) and US$ 0.20 (Improved menus) per child and day (Table 3). The CO_2_eq emissions and water footprints of the baseline school food baskets were 165g/46L (Base menus) and 367g/74L (Improved menus) per meal (Table 3).

**Table 3.** Changes in cost and environmental impacts, as well as the average relative deviation for each applied model.

|  | Baseline<br>cost | Optimised<br>cost | Baseline<br>CO<sub>2</sub>eq | Change<br>in<br>CO<sub>2</sub>eq | Baseline<br>water<br>use | Change<br>in<br>water<br>use | ARD |
| --- | --- | --- | --- | --- | --- | --- | --- |
|  | USD | USD | g | % | L | % | % |
| Model 1<sup>a</sup> | 0.10 | 0.12 | 165 | 98 | 46 | -37 | 1528 |
| Model 2<sup>b</sup> | 0.10 | 0.10 | 165 | 33 | 46 | -14 | 1272 |
| Model 3<sup>a</sup> | 0.20 | 0.10 | 367 | 347 | 74 | 232 | 589 |
| Model 4<sup>a</sup> | 0.20 | 0.13 | 367 | 213 | 74 | 101 | 384 |
| Model 1<sup>c</sup> | 0.10 | 0.12 | 165 | 43 | 46 | -37 | 429 |
| Model 3<sup>c</sup> | 0.20 | 0.10 | 367 | 306 | 74 | 157 | 316 |
| Model 4<sup>c</sup> | 0.20 | 0.13 | 367 | 248 | 74 | 113 | 182 |
| Model 5<sup>d</sup> | 0.20 | 0.99 | 367 | 183 | 74 | 157 | 127 |

<sup>a</sup>Model with cost, nutrient and food-based constraints.
<sup>b</sup>Model with cost and nutrient constraints only (a feasible solution could not be achieved when including either food-based or acceptability constraints, hence results are only available for one run of Model 2).
<sup>c</sup>Model with cost, nutrient and acceptability constraints.
<sup>d</sup>Model without a cost constraint but with nutrient, food-based, and acceptability constraints.
CO<sub>2</sub>eq = carbon dioxide equivalents.
ARD = average relative deviation.

### Optimized school food baskets

It was possible to meet cost and nutrient targets in all optimised models (Tables 2 and 3). Active constraints that prevented a higher similarity to baseline school food baskets were energy, zinc, and vitamins A, B12, and C (Table 2). When these constraints were met, the demands on the provision of protein, iron, and folate were automatically achieved, meaning that these nutrients never turn critical if the supply of energy, zinc, and vitamins A, B12, and C is sufficient. Food-based constraints were only achievable when the acceptability constraint of keeping at least 20% of the originally consumed foods was not applied (with the exception of Model 5 without a budget constraint). In one case, i.e. the optimisation of the Base menus, considering both operational costs and inflation (Model 2), a feasible solution was only found when both food-based and acceptability constraints were not applied, hence resulting in a diet with no animal foods, no fats, oils, and grains (Supplementary Figure 1, Table 4). When applying only nutrient and food-based constraints, amounts of single foods ranged between 4.4 to 16.3-times higher compared to the baseline food baskets (no data shown), with the highest ARD being for Model 1 (Table 3), indicating a very high deviation from baseline. When implementing acceptability constraints, but excluding food-based constraints, the ARDs were lower, ranging between 127-350%, with the lowest ARD (127%) in Model 5. In this case, (based on the model using improved recipes and considering both operational costs and inflation but without a budget constraint), meals would cost nearly US$ 1 and meet nutrient, food-based, and acceptability constraints. Overall, the ARDs were higher for the financially more restricted models, making them less culturally acceptable. For example, increasing the budget from US$ 0.12 in Model 3 to US$ 0.16 in Model 4 (resulting in cost constraints of US$ 0.10 and US$ 0.13 for Models 3 and 4, respectively, when considering operational costs and inflation), nearly halved the ARD from 350 to 175%.

**Table 4.** Absolute quantities of food groups at baseline and in the optimized models.

|  | Baseline<br>Base<br>menus | Model<br>1<sup>a</sup> | Model<br>1<sup>b</sup> | Model<br>2<sup>c</sup> | Baseline<br>Improved<br>menus | Model<br>3<sup>a</sup> | Model<br>3<sup>b</sup> | Model<br>4<sup>a</sup> | Model<br>4<sup>b</sup> | Model<br>5<sup>c</sup> |
| --- | --- | --- | --- | --- | --- | --- | --- | --- | --- | --- |
| Fats and oils | 2.8 | 0.0 | 0.7 | 0.0 | 8.8 | 0.0 | 1.8 | 0.2 | 3.3 | 8.8 |
| Animal foods | 1.0 | 20.0 | 0.6 | 0.0 | 9.4 | 234.0 | 242.8 | 212.8 | 220.7 | 59.9 |
| Fruits and<br>vegetables | 43.6 | 133.0 | 67.3 | 37.1 | 23.6 | 133.0 | 44.3 | 133.0 | 19.2 | 133.0 |
| Grains | 31.3 | 0.0 | 23.5 | 0.0 | 40.0 | 0.0 | 30.0 | 0.0 | 30.0 | 40.0 |
| Legumes, nuts,<br>seeds | 7.5 | 30.0 | 22.1 | 61.1 | 14.5 | 30.0 | 2.9 | 30.0 | 7.7 | 30.0 |
| Roots and tubers | 6.7 | 24.8 | 41.7 | 76.7 | 6.7 | 0.0 | 1.3 | 2.8 | 3.8 | 6.7 |

<sup>a</sup>Model with cost, nutrient and food-based constraints.
<sup>b</sup>Model with cost, nutrient and acceptability constraints.
<sup>c</sup>Model with cost and nutrient constraints only (a feasible solution could not be achieved when including either food- based or acceptability constraints, hence results are only available for one run of Model 2).
<sup>d</sup>Model without a cost constraint but with nutrient, food-based, and acceptability constraints.

In general, favoring food-based targets over cultural acceptability within a constrained budget meant the exclusion of entire food groups and large increases in others (Figure 1, Table 4). For example, all models excluding acceptability constraints completely/nearly eliminated Fats and oils and Grains, and in two cases (Models 3 and 4), Roots and tubers, given that there were no explicit food targets defined for such groups. At the same time, large increases were seen in other food groups such as animal foods that increased 20-25 fold in models 1, 3 and 4.

**Figure 1.**
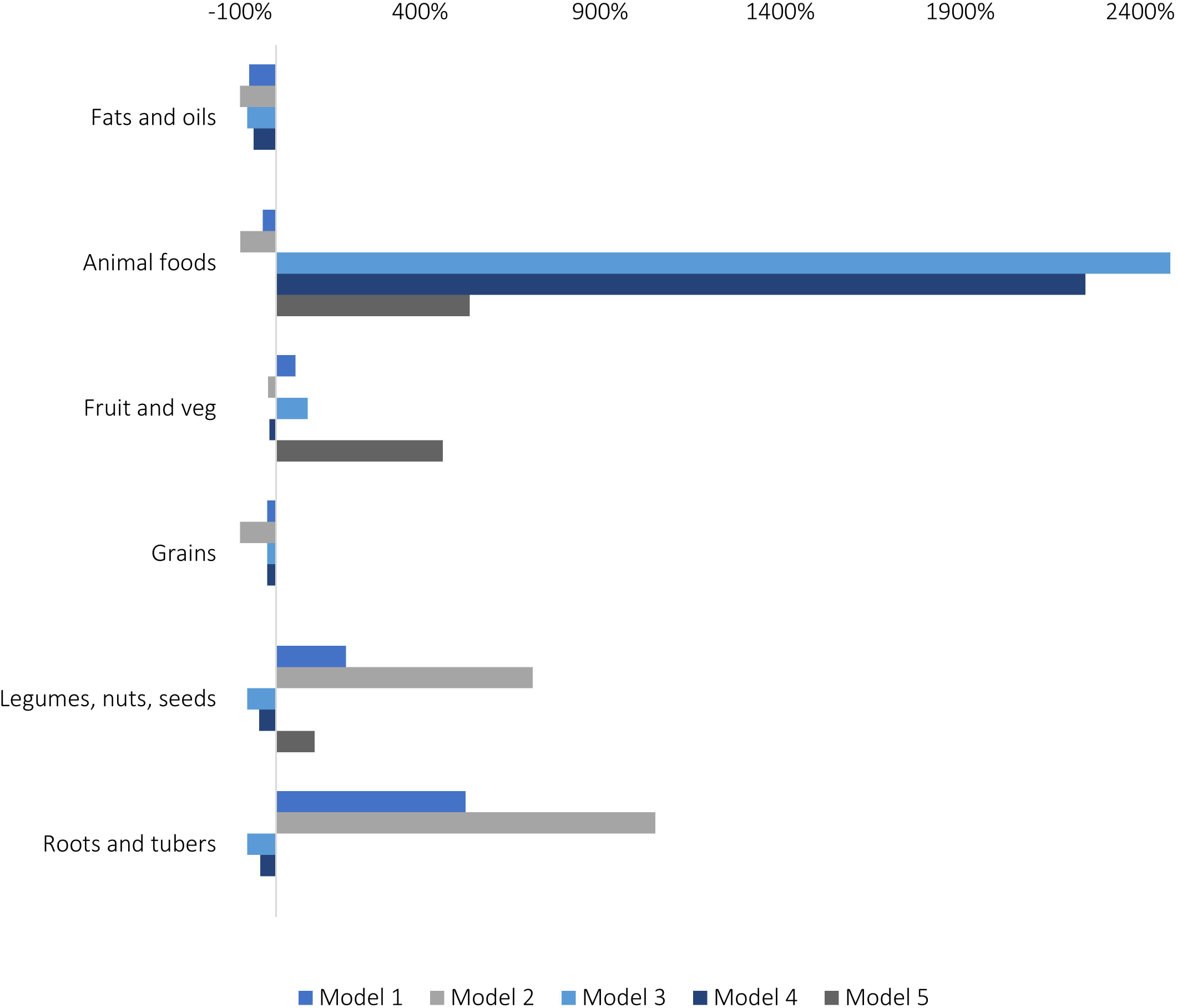
Changes in food groups in models 1-5 including acceptability and not food-based constraints. Models 1, 3 and 4 include nutrient, cost and acceptability constraints. Model 2 includes only nutrient and cost constraints. Model 5 includes nutrient, cost, food-based and acceptability constraints.

When favoring acceptability constraints over food-based constraints, no food group was eliminated and the changes to food groups are more balanced (except for animal foods that increase considerably in most models) (Figure 1, Table 4). The model without a budget constraint, and with both acceptability and food-based constraints included (Model 5) was the most conservative with regards to changes in food groups (Figure 1, Table 4). Here, Fats and oils, Grains, and Roots and tubers remained completely unchanged, and the remaining food groups only increased by ∼100- 500% compared to baseline school food baskets (Figure 1).

Compared to the baseline school food baskets, GHGE were increased in almost all models (up to +347% in Model 3 with only FBDG and no acceptability constraints) (Table 3) due to the abovementioned increase in animal foods in the optimized school food baskets compared to baseline (Table 4). Similarly, water use was increased in all models but Models 1 and 2 (Table 3).

## Discussion

### Main findings

In this study, linear programming was demonstrated to be capable of guiding the development of more nutritionally adequate and cost-effective meals for school-aged children in Ghana. Our findings show that baseline school food baskets were significantly deficient in energy, protein, and several critical micronutrients, with substantial disparities from nutrient targets. This deficiency is grounded on the fact that the children receive too little food with inadequate micronutrient density. The optimized diets, derived from our model, addressed these deficiencies while at the same time adhering to defined cost constraints, ranging between US$ 0.10 and US$ 0.13. However, when meeting these cost benchmarks, the optimized school food baskets exhibited considerable deviations from baseline amounts, indicating that achieving nutrient adequacy with a limited budget may require pronounced changes to established school meal provisioning. Moreover, it was noted that food-based constraints could only be met effectively when acceptability constraints, applied to minimize the deviation of individual foods from baseline school food supplies, were excluded, leading to school food baskets omitting entire food groups. The school food baskets examined in Ghanaian schools were found to consist of low nutrient density foods that are not able provide an adequate proportion of nutrients necessary for healthy growth and development in children. The dramatic changes necessary to achieve nutritional adequacy are caused by the fact that (expensive) foods, particularly of animal origin, are almost absent from the observed school meal food basket. Adding these foods to school meal menus is necessary to provide key micronutrients such as vitamin B12, iron, and zinc. However, meeting nutritional constraints with as little change as possible to the baseline food basket of the Improved menus would cost almost US$ 1 per meal, almost ten times more than the currently available budget when considering operational costs and inflation, highlighting important trade-offs between program financing, food quality and coverage.

The increased reliance on animal products in the optimized school food baskets contributed to a rise in GHGE, with emissions rising by as much as 347% compared to baseline school food baskets, surpassing planetary boundaries for climate change (31). Our findings thus highlight the challenge of balancing cost and nutritional needs with consumer preferences and climate targets in the context of limited food diversity and global challenges such as rising food prices.

### Interpretation

The baseline school food basket for the Base (current) menus was deficient in multiple nutrients, including iron, zinc, vitamin A, folate and vitamin B12. This mirrors current deficiencies among Ghanaian children, which include iron, vitamin A, and iodine (32,33). Iron deficiency anemia is particularly prevalent, affecting 12.2% of children under five (32). Vitamin A deficiency affects 20.8% of children, while iodine deficiency is also common (32,33).

Previous optimization research in Ghana (14) also illustrated the difficulty in achieving nutritional adequacy under financial constraints while considering cultural acceptability. As with the school meals in the study at hand, vitamins A, B12, and C also appeared as active constraints in highly affordable diets for Ghanaian families (14). However, the nutritionally optimized school food baskets for Ghanaian families deviated considerably less from the baseline consumption compared to the ARD values found in this study, primarily due to the much larger number of foods (N= 152). Therefore, expanding the number of foods being used for Ghanaian school menus (N=45) with those available at Ghanaian markets may achieve nutritionally adequate menus at lower cost than those found in the current study. The foods selected for this menu improvement could be chosen particularly with respect to cost and nutrient density for nutrients that appear as active constraints (zinc and the vitamins A, B12, and C). One example of such a food is red palm oil, an affordable resource for vitamin A (34).

In our study, food-based constraints were only successfully met when acceptability constraints were removed, which resulted in the exclusion of entire food groups from the school meal provisions.

This is grounded on suggested goals of implementing minimum amounts (rather than ranges) of fruits, vegetables, pulses, and animal-source foods (ASF). These goals deviate considerably from the reality of Ghanaian school menus, explaining the difficulty in achieving optimized solutions for nutritional adequacy which closely resemble what is currently provided. This applies particularly to animal-source foods, which can significantly improve growth, cognitive function, and academic performance (35,36). The benefits of ASF are presumably attributed to their high content of bioavailable micronutrients, including iron, zinc, vitamin B12, and vitamin A (37), which also appeared as active constraints in the optimized models of this study. However, access to animal-source foods remains limited in many LMICs, and more research is needed to determine optimal intake levels and strategies for increasing animal-source foods consumption among vulnerable populations (38).

The optimisation of school meals in Ghana, while adhering to nutritional and cost constraints, resulted in an increase of animal products and GHGE in nearly all models applied. This stands in contrast to similar optimization of diets/school meals in high-income countries (HICs) such as Sweden (15,39,40), the UK (41), and Germany (42), where nutritionally-adequate and sustainable diets were achieved by a reduction of animal product consumption. This discrepancy is understandable, given that baseline diets in these HICs typically are much more expensive, containing a considerably higher proportion of animal products. Furthermore, diets in HICs typically exhibit a large overall food diversity. This diversity allows for nutritional inadequacies to be addressed much more easily, compared with the limited spectrum of foods used for school meals in Ghana. Food diversity thus seems to bear important weight when determining model outcomes. Supporting this perspective, research from Mozambique (43) found that increasing food diversity would increase the cost 3-fold compared to a minimum cost diet. The same study showed that optimizing food baskets using local, nutrient-dense foods could meet nutrient recommendations. It also emphasized the higher cost associated with achieving a fully nutritious basket which also remains culturally acceptable. Our model without a cost constraint (Model 5) aligns with these findings, being the model with the lowest deviation from baseline, but with the highest cost.

### Strengths and limitations

Our study employed a robust optimization framework that aimed to balance nutritional adequacy with cost and cultural acceptability constraints, allowing for a detailed analysis of how different scenario for school meals in Ghana could be designed. It combines an array of plausible scenarios for designing improved school meals in Ghana within limited budgets, based on real local data on school food baskets. By explicitly examining food-based constraints, the study highlights the importance of integrating more diverse food groups into meal planning, thus ensuring that dietary? diversity is addressed in the formulations.

Our models are limited by the fact that they only operate based on the foods that were available from the collected data/selected recipes. Achieving full nutritional adequacy for Ghanaian school meals, potentially other nutrients and foods should also be considered. For example, iodine deficiency is a persistent health problem for children in West Africa (32). Future optimisation studies could investigate what other foods could be available at local level, through home-grown school feeding (HGSF), and explore the potential of incorporating such foods into the optimization models. Future optimisation studies could also investigate what other foods could be available at local level that could be promoted through HGSF. For example, African indigenous leafy vegetables, have been associated with increased dietary diversity and improved micronutrient levels, including zinc, iron, and vitamin A (44,45).

The necessity to exclude acceptability constraints (in this case aiming to reflect both local food preferences and what the cooks prepare which is dictated by budget constraints) in order to meet food-based targets may limit the practical applicability of the optimized school food baskets. This could lead to scenarios that are not feasible in real-world settings where local conditions (e.g. budgets available to provide school meals) and preferences play a significant role. The exclusion of entire food groups— in the absence of explicit targets for such groups— when acceptability constraints are removed also raises concerns about the nutritional quality of meals, which may arise from deficient supply of nutrients not considered in the current optimizations. This could lead to diets that, while optimized for cost and some nutrient targets, may still be deficient in other important vitamins, minerals, or macronutrients such as iodine, vitamin E, and poly-unsaturated (omega-3 and omega-6) fatty acids. Furthermore, the specific context of the study may limit the generalizability of the findings to other regions or settings that have different cultural, economic, and dietary practices. Without a diverse array of contexts being tested, the conclusions may not apply universally.

While optimization provides a useful approach to create food baskets that consider multiple priorities at the same time, this approach requires advanced technical expertise and significant time investment, limiting scalability in the context of meal planning. For meal planning, there is a need for more user-friendly solutions that cater to the needs of cooks and caterers responsible for delivering and preparing school meals in practice in countries like Ghana.

### Conclusions

In summary, this study highlights the utility of linear programming as a powerful tool for optimizing school food baskets in Ghana, and for understanding the complex synergies and trade-offs between nutritional adequacy, cost-effectiveness, cultural acceptability, and environmental sustainability. The results reveal significant deficiencies in current school meal menus in Ghana, indicating an urgent need for improvement in the service delivery. Furthermore, our findings indicate that current inflationary trends are likely to exacerbate the challenge of meeting targets for nutrients, food groups, and palatability with existing budget allocations. Current budgets will likely need to be increased up to 6-fold to meet all requirements for nutrients, food groups, and feasibility/cultural acceptability. Though current environmental bounds for the program are low, GHGE and water use projections for improved menus will require attention as the program service improves. Important gaps remain in terms of providing guidance on how to operationalize these findings, including in the short-term with current funding constraints. On the medium term, evidence from this study suggests that Government planners and development partners increase the financial investment in the GSFP to enable the program to achieve its ultimate goal of creating a healthier, more equitable food environment for children in Ghana, whilst also recognizing that addressing local realities, governance and related constraints will be crucial for the success and sustainability of programs.

## Data Availability

All data produced in the present study are available upon reasonable request to the authors.

## Abbreviations

ARD: Average relative deviation
ASF: Animal-source foods
CO2eq: Carbon dioxide equivalents
FAO: Food and Agriculture Organization of the United Nations
GHGE: Greenhouse gas emissions
GSFP: Ghana School Feeding Program
HGSF: Home-grown school feeding
HICs: High-income countries
LP: Linear programming
RD: Relative deviation
SoP: Standard Operating Procedures
TRD: Total relative deviation
WU: Water use

## Notes

### Competing Interest Statement

The authors have declared no competing interest.

### Funding Statement

This study did not receive any funding.

## References

1. The State of Food Security and Nutrition in the World 2024 [Internet]. FAO; IFAD; UNICEF; WFP; WHO; 2024 [cited 2025 Mar 27]. Available from: https://openknowledge.fao.org/handle/20.500.14283/cd1254en

2. Norris SA, Frongillo EA, Black MM, et al. Nutrition in adolescent growth and development. The Lancet. 2022 Jan;399(10320):172–84.

3. Bailey RL, West KP, Black RE. The epidemiology of global micronutrient deficiencies. Ann Nutr Metab. 2015;66 Suppl 2:22–33.

4. Black RE, Victora CG, Walker SP, et al. Maternal and child undernutrition and overweight in low-income and middle-income countries. The Lancet. 2013 Aug 3;382(9890):427–51.

5. WFP. State of School Feeding Worldwide 2022. Rome: World Food Programme; 2022.

6. Alderman H, Bundy D, Gelli A. School Meals Are Evolving: Has the Evidence Kept Up? World Bank Res Obs. 2024 Jul 18;39(2):159–76.

7. Global Nutrition Report | Country Nutrition Profiles - Global Nutrition Report [Internet]. [cited 2024 Nov 20]. Available from: https://globalnutritionreport.org/resources/nutrition-profiles/africa/western-africa/ghana/

8. The World Bank. International development association project appraisal document on a proposed credit in the amount of us$100.0 million to the republic of Ghana for the Ghana productive safety net project 2. Washington DC: The World Bank; 2021.

9. Aurino E, Gelli A, Adamba C, et al. Food for Thought?: Experimental Evidence on the Learning Impacts of a Large-Scale School Feeding Program. J Hum Resour. 2023 Jan;58(1):74–111.

10. Gelli A, Aurino E, Folson G, et al. A School Meals Program Implemented at Scale in Ghana Increases Height-for-Age during Midchildhood in Girls and in Children from Poor Households: A Cluster Randomized Trial. J Nutr. 2019 Aug;149(8):1434–42.

11. Partnership for Child Development, Department of Infectious Disease Epidemiology, Imperial College, London, UK, Parish A, Gelli A, et al. Trade-offs in costs, diet quality and regional diversity: an analysis of the nutritional value of school meals in Ghana. Afr J Food Agric Nutr Dev. 2015 Oct 2;15(71):10217–40.

12. Owusu JS, Colecraft EK, Aryeetey RNO, et al. Comparison of Two School Feeding Programmes in Ghana, West Africa. Int J Child Health Nutr. 2016 Sep 2;5(2):56–62.

13. De Carvalho F, Dom BS, Fiadzigbey MM, et al. Ghana School Feeding Program: re-tooling for a sustainable future. Partnership for Child Development and the Ministry of Local Government and Rural Development, Ghana; 2011.

14. Nykänen EPA, Dunning HE, Aryeetey RNO, et al. Nutritionally Optimized, Culturally Acceptable, Cost-Minimized Diets for Low Income Ghanaian Families Using Linear Programming. Nutrients. 2018 Apr 7;10(4):461.

15. Eustachio Colombo P, Patterson E, Lindroos AK, et al. Sustainable and acceptable school meals through optimization analysis: an intervention study. Nutr J. 2020 Dec;19(1):61.

16. Elinder LS, Eustachio Colombo P, Patterson E, et al. Successful Implementation of Climate-Friendly, Nutritious, and Acceptable School Meals in Practice: The OPTIMAT^TM^ Intervention Study. Sustainability. 2020 Oct 14;12(20):8475.

17. André E, Eustachio Colombo P, Schäfer Elinder L, et al. Acceptance of Low-Carbon School Meals with and without Information—A Controlled Intervention Study. J Consum Policy. 2024 Mar;47(1):109–25.

18. Darmon N, Ferguson EL, Briend A. A Cost Constraint Alone Has Adverse Effects on Food Selection and Nutrient Density: An Analysis of Human Diets by Linear Programming. J Nutr. 2002 Jan 12;132(12):3764–71.

19. FAO. School Meal Nutrition Guidelines and Standards: Exploring challenges, finding solutions and opportunities for institutionalization - Stakeholder Consultation Workshop Report. Rome: FAO; Unpublished.

20. Guidance for the Use of Standard and Non-Standard Recipes in Quantitative 24-Hour Dietary Recall Surveys: The Simple Ingredient Method | USAID Advancing Nutrition [Internet]. [cited 2025 Mar 27]. Available from: https://www.advancingnutrition.org/resources/guidance-use-standard-and-non-standard-recipes-quantitative-24-hour-dietary-recall

21. School Meal Planner (SMP) PLUS | WFP Innovation [Internet]. [cited 2025 Mar 17]. Available from: https://innovation.wfp.org/project/school-meal-planner-smp-plus

22. Vincent A, Grande F, Compaoré E, et al. Food Composition Table for Western Africa (2019) User Guide & Condensed Food Composition Table / Table de composition des aliments FAO/INFOODS pour l’Afrique de l’Ouest (2019) Guide d’utilisation & table de composition des aliments condensée. Rome: FAO; 2019.

23. Poore J, Nemecek T. Reducing food’s environmental impacts through producers and consumers. Science. 2018 Jun;360(6392):987–92.

24. Gazan R, Brouzes CMC, Vieux F, et al. Mathematical Optimization to Explore Tomorrow’s Sustainable Diets: A Narrative Review. Adv Nutr. 2018 Sep 1;9(5):602–16.

25. Parlesak A, Tetens I, Dejgard Jensen J, et al. Use of Linear Programming to Develop Cost-Minimized Nutritionally Adequate Health Promoting Food Baskets. PloS One. 2016;11(10):e0163411.

26. Dantzig GB. 1947. Maximization of a linear function of variables subject to linear inequality. In: Koopmans TC, editor Activity analysis of production and allocation. New York-London: Wiley & Chapman-Hall; 1951. p. 339–47.

27. Nocedal J, Wright SJ. Numerical optimization. New York: Springer; 2006.

28. Mason AJ. OpenSolver - An Open Source Add-in to Solve Linear and Integer Progammes in Excel. In: Klatte D, Lüthi HJ, Schmedders K, editors. Operations Research Proceedings 2011. Berlin, Heidelberg: Springer Berlin Heidelberg; 2012. p. 401–6.

29. Eustachio Colombo P, Patterson E, Elinder LS, et al. Optimizing School Food Supply: Integrating Environmental, Health, Economic, and Cultural Dimensions of Diet Sustainability with Linear Programming. Int J Environ Res Public Health. 2019 Jan;16(17):3019.

30. FAO. FAO and WFP (forthcoming). Developing holistic nutrition guidelines and standards for school meals: a global methodology. Rome: FAO;

31. World Wildlife Fund. One Planet Plate 2019 – kriterier och bakgrund (One Planet Plate 2019 – criteria and background) [Internet]. 2019 [cited 2020 Jan 30]. Available from: https://wwwwwfse.cdn.triggerfish.cloud/uploads/2019/04/kriterier-fr-one-planet-plate-rev-2019.pdf

32. Wegmüller R, Bentil H, Wirth JP, et al. Anemia, micronutrient deficiencies, malaria, hemoglobinopathies and malnutrition in young children and non-pregnant women in Ghana: Findings from a national survey. Gebremedhin S, editor. PLOS ONE. 2020 Jan 30;15(1):e0228258.

33. Egbi G. OF IODINE DEFICIENCY AND ANAEMIA AMONG 2-10 YEAR- OLD GHANAIAN CHILDREN. In 2012 [cited 2025 Mar 5]. Available from: https://www.semanticscholar.org/paper/OF-IODINE-DEFICIENCY-AND-ANAEMIA-AMONG-2-10-YEAR-Egbi/3fcec63833acb9403a86002d5881977954fae306

34. Rice AL, Burns JB. Moving from Efficacy to Effectiveness: Red Palm Oil’s Role in Preventing Vitamin A Deficiency. J Am Coll Nutr. 2010 Jun;29(sup3):302S–313S.

35. Grillenberger M, Neumann CG, Murphy SP, et al. Intake of micronutrients high in animal-source foods is associated with better growth in rural Kenyan school children. Br J Nutr. 2006 Feb;95(2):379–90.

36. Khonje MG, Qaim M. Animal-sourced foods improve child nutrition in Africa. Proc Natl Acad Sci. 2024 Dec 10;121(50):e2319009121.

37. Beal T. Achieving dietary micronutrient adequacy in a finite world. One Earth. 2021 Sep 17;4(9):1197– 200.

38. Eaton JC, Rothpletz-Puglia P, Dreker MR, et al. Effectiveness of provision of animal-source foods for supporting optimal growth and development in children 6 to 59 months of age. Cochrane Developmental, Psychosocial and Learning Problems Group, editor. Cochrane Database Syst Rev [Internet]. 2017 Oct 3 [cited 2025 Jun 6]; Available from: https://doi.wiley.com/10.1002/14651858.CD012818

39. Eustachio Colombo P, Elinder LS, Lindroos AK, et al. Designing Nutritionally Adequate and Climate-Friendly Diets for Omnivorous, Pescatarian, Vegetarian and Vegan Adolescents in Sweden Using Linear Optimization. Nutrients. 2021 Jul 22;13(8):2507.

40. Eustachio Colombo P, Elinder LS, Nykänen EPA, et al. Developing a novel optimisation approach for keeping heterogeneous diets healthy and within planetary boundaries for climate change. Eur J Clin Nutr. 2024 Mar;78(3):193–201.

41. Milner J, Green R, Dangour AD, et al. Health effects of adopting low greenhouse gas emission diets in the UK. BMJ Open. 2015 Apr 30;5(4):e007364–e007364.

42. Masino T, Colombo PE, Reis K, et al. Climate-friendly, health-promoting, and culturally acceptable diets for German adult omnivores, pescatarians, vegetarians, and vegans – a linear programming approach. Nutrition. 2023 May;109:111977.

43. Parlesak A, Geelhoed D, Robertson A. Toward the Prevention of Childhood Undernutrition: Diet Diversity Strategies Using Locally Produced Food Can Overcome Gaps in Nutrient Supply. Food Nutr Bull. 2014 Jun 1;35(2):191–9.

44. John Wakhanu, Hudson Nyambaka, Judith Kimiywe, et al. Consumption of African Indigenous Vegetables Improves Children’s body Fat Free Mass in Machakos County, Kenya. J Food Sci Eng [Internet]. 2019 Aug 28 [cited 2025 Mar 5];9(8). Available from: http://www.davidpublisher.org/index.php/Home/Article/index?id=41838.html

45. Department of Community Health and Epidemiology in the School of Public Health and Applied Human Sciences, Kenyatta University, Kenya., Lubeka C, Kimiywe J, et al. Impact of school garden on dietary diversity and micronutrient level of pre-school children in Makueni County –Kenya. Res J Food Sci Nutr. 2020 Oct 30;5(4):85–97.

