## Supplementary information for "Optimizing school meals in Ghana: integrating new food and nutrient standards with aspects of affordability, cultural acceptability and environmental sustainability"

### Materials and methods

#### Dishes used for the Base menus and example of recipes

**Dish name**

Banku and Tomato Stew

Banku and Okro Soup

Tuo Zaafi & Green Leafy Vegetables

Banku and Groundnut Soup

Banku and Okro Stew

Banku and Tomato Stew

Banku and Bra Leaf Soup

Banku and Palmnut Soup

Banku and Fante Fante Soup

Gari and Beans Stew

Gari and Beans

Yoro Yoro Yori Yori

Gari Balls and Okro Soup

Plain Rice and Beans Stew

Braised Rice and Tomato Stew

Shinkafa Tomato Stew

Waakye Jollof

Stewed Waakye

**Banku and Bra Leaf Soup (Base recipe)**

| **Food** | **Grams** |
| --- | --- |
| Cassava dough | 15 |
| Corn dough | 45 |
| Oil palm, fruit, raw | 38 |
| Onion, fresh, raw | 0.4 |
| Beef, corned, canned | 0.09 |
| Pepper, sweet, green, fresh, raw | 0.5 |
| Fish, whole, dried | 0.1 |
| Okra, fruit, fresh, raw | 0.5 |

**Banku and Fante Fante Soup (Base recipe)**

| **Food** | **Grams** |
| --- | --- |
| Cassava dough | 41.1 |
| Corn dough | 17 |
| Salt | 0.5 |
| Palm oil, red | 2.5 |
| Onion, fresh, raw | 0.1 |
| Pepper, chilli, fresh, raw | 0.2 |
| Chilli pepper, dried | 0.2 |
| Ginger, root, raw | 0.05 |
| Garlic, flesh, raw | 0.05 |
| Tomato, red, ripe, raw | 1.5 |
| Wheat flour, white, unfortified | 1 |
| Crab, flesh (body and claw), boiled* (as part of a recipe) | 3.1 |
| Salt | 0.2 |
| Beef, corned, canned | 0.9 |
| Fish, powder | 0.8 |

**Gari and Beans (Base recipe)**

| **Food** | **Grams** |
| --- | --- |
| Cassava, grated, from fermented white cassava, toasted without oil (white gari) | 29 |
| Bean, white, dry, raw | 34.5 |
| Salt | 0.7 |
| Palm oil, red | 14.8 |
| Onion, fresh, raw | 0.3 |
| Chilli pepper, dried | 0.1 |
| Salt | 0.7 |

**Braised Rice and Tomato Stew (Base recipe)**

| **Food** | **Grams** |
| --- | --- |
| Vegetable oil, fortified with vitamin A | 0.5 |
| Onion, fresh, raw | 0.03 |
| Rice, white, polished, raw | 52.8 |
| Salt | 1 |
| Vegetable oil, fortified with vitamin A | 3.2 |
| Onion, fresh, raw | 0.4 |
| Chilli pepper, dried | 0.2 |
| Garlic, flesh, raw | 0.1 |
| Tomato, paste, concentrated, without salt | 7.6 |
| Mackerel, fillet, grilled* (without salt or fat) | 1.5 |
| Salt | 0.2 |

#### Dishes used for the Improved menus and example of recipes

**Dish name**

Banku and Tomato Stew

Banku and Okro Soup

Banku and Okro Stew

Tuo Zaafi & Green Leafy Vegetables

Banku and Groundnut Soup

Banku and Okro Stew

Banku and Tomato Stew

Banku and Bra Leaf Soup

Banku and Palmnut Soup

Banku and Fante Fante Soup

Gari and Beans Stew

Gari and Beans

Yoro Yoro Yori Yori

Gari Balls and Okro Soup

Plain Rice and Beans Stew

Jollof

Plain Rice and Tomato Stew

Braised Rice and Tomato Stew

Tuo-Zafi and Dry Okro Soup

Tuo Zaafi and Bra Leaf Soup

Waakye and Tomato Stew

Shinkafa Tomato Stew

Stewed Waakye Waakye Jollof

**Banku and Bra Leaf Soup (Improved recipe)**

| **Food** | **Grams** |
| --- | --- |
| Cassava dough | 40 |
| Corn dough | 75 |
| Soya bean, dry, raw | 20 |
| Oil palm, fruit, raw | 20 |
| Native eggplant, fruit, raw (turkey berries) | 5 |
| Onion, fresh, raw | 5 |
| Pepper, chilli, fresh, raw | 5 |
| Chilli pepper, dried | 5 |
| Ginger, root, raw | 5 |
| Garlic, flesh, raw | 5 |
| Tomato, red, ripe, raw | 10 |
| Groundnut paste, from groundnuts only | 10 |
| Sweet potato, orange flesh, raw | 20 |
| Fish, powder | 15 |
| Texturized soya protein (TSP) | 20 |

**Banku and Fante Fante Soup (Improved recipe)**

| **Food** | **Grams** |
| --- | --- |
| Cassava dough | 0 |
| Corn dough | 0 |
| Soya bean, dry, raw | 0 |
| Palm oil, red | 10 |
| Onion, fresh, raw | 5 |
| Pepper, chilli, fresh, raw | 5 |
| Chilli pepper, dried | 5 |
| Ginger, root, raw | 5 |
| Garlic, flesh, raw | 5 |
| Tomato, red, ripe, raw | 10 |
| Wheat flour, white, unfortified | 20 |
| Sweet potato, orange flesh, raw | 10 |
| Texturized soya protein (TSP) | 0 |
| Fish, powder | 0 |
| Fish, tuna, smoked | 9 |

**Gari and Beans (Improved recipe)**

| **Food** | **Grams** |
| --- | --- |
| Bean, white, dry, raw | 60 |
| Palm oil, red | 20 |
| Onion, fresh, raw | 5 |
| Pepper, chilli, fresh, raw | 5 |
| Ginger, root, raw | 5 |
| Garlic, flesh, raw | 5 |
| Fish, powder | 15 |
| Cassava, grated, from fermented white cassava, toasted without oil (white gari) | 20 |

**Braised Rice and Tomato Stew (Improved recipe)**

| **Food** | **Grams** |
| --- | --- |
| Vegetable oil, fortified with vitamin A | 10 |
| Palm oil, red | 5 |
| Onion, fresh, raw | 5 |
| Chilli pepper, dried | 5 |
| Pepper, chilli, fresh, raw | 5 |
| Ginger, root, raw | 5 |
| Garlic, flesh, raw | 5 |
| Tomato, red, ripe, raw | 10 |
| Tomato, paste, concentrated, without salt | 20 |
| Fish, powder | 0 |
| Egg, boiled | 0 |
| Fish, tuna, smoked | 9 |

### Results

|  | Model 1 | Model 2 | Model 3 | Model 4 | Model 5 |
| --- | --- | --- | --- | --- | --- |
| Fats and oils | -100% | -100% | -100% | -97.7051% | -6.425e-07% |
| Animal foods | 1872.71% | -99.1649% | 2389.56% | 2164.16% | 537.74% |
| Fruit and veg | 204.899% | -22.5761% | 463.533% | 463.533% | 463.533% |
| Grains | -99.9346% | -99.9346% | -100% | -100% | -1.2355e-06% |
| Legumes, nuts, seeds | 299.47% | 713.211% | 106.838% | 106.838% | 106.838% |
| Roots and tubers | 273.391% | 1053.46% | -100% | -58.6207% | -1.799e-07% |

#### **Supplementary Figure 1.** Changes in food groups in models 1-5 including food-based and not acceptability constraints. Models 1, 3 and 4 include nutrient, cost and food-based constraints. Model 2 includes only nutrient and cost constraints. Model 5 includes nutrient, cost, food-based and acceptability constraints.
